# Aging, Structural, and Neighborhood-Environmental Correlates of Cardiovascular–Kidney–Metabolic Risk Factor Burden Across U.S. Census Tracts

**DOI:** 10.64898/2026.11.12.26367979

**Authors:** Ahmed Batobara, Maha Alfaraidhy, Bekir Tanriover, Yiwen Liu, Abdulla A. Damluji, Amanda R. Vest, Chris Lim

## Abstract

**Background:** The American Heart Association defined Cardiovascular–Kidney–Metabolic (CKM) syndrome in 2023. Census-tract studies often examined one condition without spatial modeling. It is unclear where combined Stage 1–2 burden concentrates and how it relates to aging, environment, and socioeconomic conditions.

**Methods:** We built a CKM burden index for 70,336 tracts from five CDC PLACES crude prevalences (obesity, diabetes, hypertension, hypercholesterolemia, chronic kidney disease) and compared it with NHANES 2017–2020. In 69,530 contiguous-U.S. tracts, we fit Bayesian spatial (BYM2) models with demographic, socioeconomic, and environmental predictors and aging × environment interactions, plus descriptive machine learning. The primary model excluded behavioral measures sharing the outcome’s BRFSS source; a second added them. Credible intervals (CrIs) are conditional on PLACES estimates.

**Results:** Agreement with NHANES was modest. Burden clustered spatially (Moran’s I = 0.657) and highest in the South. Behavioral measures had the largest machine-learning importance (52.6%), partly from shared sourcing. Each percentage-point increase in adults ≥65 years was associated with 0.112 units higher burden (95% CrI 0.111–0.113) outside low-income, low-access (LILA) tracts. LILA tracts had 0.10–0.14 units higher burden from the 25th aging percentile but not consistently in the youngest decile. Aging and socioeconomic associations held across sensitivity analyses. The primary model’s residual Southern excess approached zero after adding behavioral measures, which may reflect shared measurement, real differences in these behaviors, or both.

**Conclusions:** CKM burden is spatially clustered and highest in the South. LILA tracts had modestly higher burden above the youngest tracts. These cross-sectional estimates support area-level surveillance and needs assessment.

**Clinical Perspective:** *What Is New?:*

- Across 69,530 US census tracts, a combined cardiovascular-kidney-metabolic burden index built from CDC PLACES was clustered in space and highest across the South.
- Each 1-percentage-point higher share of adults aged ≥65 years was associated with 0.112 units higher burden (crude prevalence, not age-standardized).

*What Are the Clinical Implications?:*

- Tract-level burden maps can help health systems and public health agencies direct cardiovascular-kidney-metabolic screening and prevention to high-burden areas, including the South, as area-level needs assessment rather than individual risk prediction.

## 1. Introduction

In October 2023, the American Heart Association issued a presidential advisory that recast the conditions of obesity, diabetes mellitus, dyslipidemia, hypertension, chronic kidney disease (CKD), and atherosclerotic cardiovascular disease into a single integrated term: Cardiovascular– Kidney–Metabolic (CKM) syndrome^1^. The advisory introduced a four-stage framework, from Stage 0 (no risk factors) through Stage 1 (excess or dysfunctional adiposity), Stage 2 (metabolic risk factors and/or moderate-to-high-risk CKD), Stage 3 (subclinical cardiovascular disease or very-high-risk CKD), and Stage 4 (clinical cardiovascular disease, including kidney failure subcategory 4b), and called for action on the social determinants of health that pattern progression through these stages. Because the intensity of monitoring and treatment escalates across these stages, identifying which populations and places carry the greatest CKM burden is central to directing screening, prevention, and management.

Population aging, structural disadvantage, and environmental exposure might shape the geography of CKM burden. For instance, adults aged ≥65 years make up about 16.8% of the U.S. population, and the prevalence of every CKM component rises steeply with age^2,3^. The unequal geography of structural disadvantage is captured by Krieger’s Index of Concentration at the Extremes (ICE)^4^, whose association with cardiometabolic outcomes persists after individual-level adjustment^5,6^. Long-term ambient air pollution is associated with cardiopulmonary mortality^7^, summarized in an AHA scientific statement^8^, and with chronic kidney disease and diabetes^9,10^; prospective evidence links pollutant mixtures to the onset and progression of cardio-renal-metabolic multimorbidity itself^11^. The nearest prior nationwide tract-level analysis compared seven composite neighborhood indices against median household income as predictors of coronary heart disease, stroke, and chronic kidney disease, with obesity, hypertension, and diabetes as secondary outcomes, and found similar associations^12^. Two of those outcomes, coronary heart disease and stroke, are Stage 4 clinical endpoints in the AHA framework. However, where the combined Stage 1–2 CKM burden that precedes them concentrates, once spatial dependence is modeled, has not been described nationally at the census-tract level. It is also unknown whether this burden is highest in tracts whose populations are older and more exposed to environmental hazards.

We construct a continuous tract-level index of the AHA CKM Stage 1–2 burden using CDC PLACES estimates of the five core cardiometabolic indicators and compare it against National Health and Nutrition Examination Survey (NHANES) 2017–2020 individual-level co-occurrence patterns. Then, we relate the index to demographic, socioeconomic, and environmental predictors in a hierarchical Bayesian BYM2 spatial regression, cross-checked by ordinary least squares with residual-autocorrelation diagnostics, and in an XGBoost/SHAP machine-learning model (§S2.4). All formal inferential claims rest on the spatial regression models; the machine-learning model is exploratory and is not used to test population-level hypotheses.

We address four aims:

1. *Construction and external comparison.* Construct a tract-level CKM burden index that summarizes the prevalence of five conditions in the AHA Stage 1–2 framework and characterize its agreement with independent individual-level NHANES estimates.
2. *Correlate decomposition.* Quantify the relative contribution of demographic, socioeconomic, and environmental predictors to the spatial distribution of CKM burden using inference from the spatial regression and exploratory decomposition from the machine-learning model.
3. *Aging × environment interaction.* Examine whether associations between neighborhood environmental characteristics and CKM burden differ according to the proportion of residents aged ≥65 years.
4. *Behavioral-block contrast.* Quantify how much of the socioeconomic-concentration signal is carried by the co-reported behavioral and healthcare-engagement block, by comparing an otherwise identical pair of spatial models fit with and without that block.

## 2. Methods

This is a cross-sectional, ecological analysis of U.S. census tracts using publicly available, de-identified secondary data; it did not constitute human-subjects research requiring institutional review board approval. Dr Batobara had full access to all the data in the study and takes responsibility for its integrity and the data analysis. Reporting follows the STROBE and RECORD guidelines (Supplemental Methods). The outcome is a continuous tract-level CKM burden index: the first principal component of five crude, non-age-standardized CDC PLACES 2022 tract prevalences (obesity, diabetes, hypertension, hypercholesterolemia, and self-reported chronic kidney disease), oriented so that higher values denote greater burden^13^. We used the 2022 release because the 2023 release has no Florida estimates for measures based on 2021 BRFSS data and the 2024 release no longer includes chronic kidney disease. Agreement with independent individual-level data was assessed against NHANES 2017–2020^14^ (§3.1).

We grouped the predictors into four blocks. Table S1a lists the source, year, spatial resolution, and units of each variable. The demographic block includes the percentage of adults aged ≥65 years and the percentage of Hispanic or Latino residents. The socioeconomic block includes Krieger’s Index of Concentration at the Extremes for race and for income, the percentage of adults with a bachelor’s degree or higher, and rurality from USDA Rural-Urban Commuting Area codes^15^. The environmental block includes air pollution (PM₂.₅ and ozone, from the EPA Downscaler), warm-season heat (mean daily maximum temperature from May to September, from PRISM^16^), walkability (EPA National Walkability Index^17^), and food access. For food access, we used the USDA flag for low-income, low-access (LILA) tracts (1-mile/10-mile definition), because it requires both low income and poor access to food. The behavioral and healthcare-engagement block includes four more PLACES measures: physical inactivity, smoking, routine checkups, and lack of health insurance. Supplemental Methods explains how we assigned the air pollution and temperature values to each tract.

Our main analysis used a hierarchical Bayesian spatial regression. Each tract had a random effect with a scaled BYM2 prior, which lets tract effects be spatially structured, unstructured, or a mix of both. Neighbors were each tract’s 8 nearest tracts (k = 8). We fit the models by integrated nested Laplace approximation (INLA) with penalized-complexity priors. Continuous predictors in interaction terms were mean-centered.

We report each association as a posterior mean with a 95% credible interval (CrI) and call it credible when the interval excludes zero. When a result combines several coefficients, such as the LILA contrast at a given age composition, we computed it from the joint posterior rather than from the separate intervals. The CrIs are conditional on the PLACES estimates; that is, they treat those estimates as fixed. They do not include the uncertainty of the PLACES model itself, whose shared county-and state-level random effects make errors correlated across tracts (§S3.7).

We fit two models on the same tracts. Model 1, the primary model, excludes the behavioral and healthcare-engagement block; Model 2 includes it. The two models answer different questions and comparing them is one of our aims. Model 1 is primary because the behavioral measures come from the same BRFSS survey cycles as the outcome, so adjusting for them may remove part of the associations we want to study. Several demographic and socioeconomic predictors are also inputs to the PLACES model (§4.5), but we keep them because they are the structure under study, whereas the behavioral measures are candidate mediators.

We used ordinary least squares, with checks for residual spatial autocorrelation, and eigenvector spatial filtering as cross-checks. We also fit a tuned XGBoost model with SHAP values, which we report descriptively only. To test the shape of the aging association, we refit Model 1 with a natural spline for % age ≥65 years (df = 4, with df = 3 and 5 as sensitivity checks; §S2.4) and compared the fits by log marginal likelihood (§3.3; Table S13). We also refit Model 1 with food-store counts from the National Neighborhood Data Archive^18^ in place of the LILA flag (§S3.10). Supplemental Methods describes the sensitivity analyses, and their results appear in Supplemental Results (Tables S8–S10 and S14) and Supplemental Discussion (Table S12). All analyses used R 4.6.0 with the sf, spdep, spmoran, R-INLA, and xgboost packages.

## 3. Results

### 3.1 Sample, index construction, and comparison with NHANES

Figure S13 shows how we built the analytic sample. Joining CDC PLACES with the American Community Survey (ACS)^19^ gave 72,335 tracts. We excluded New Jersey, whose tracts lack PLACES estimates for hypertension and high cholesterol, which left 70,336 tracts for building the CKM burden index. Then, we excluded Alaska, Hawaii, and tracts with missing covariates, which left 69,530 tracts (Table 1). We used this sample for both the OLS baseline (§3.2) and the BYM2 regression (§3.3). The first principal component (PC1) explained 76.1% of the variance in the five indicators, and all five loadings were positive and similar in size (range 0.36–0.49; Table 2).

**Table 1.**
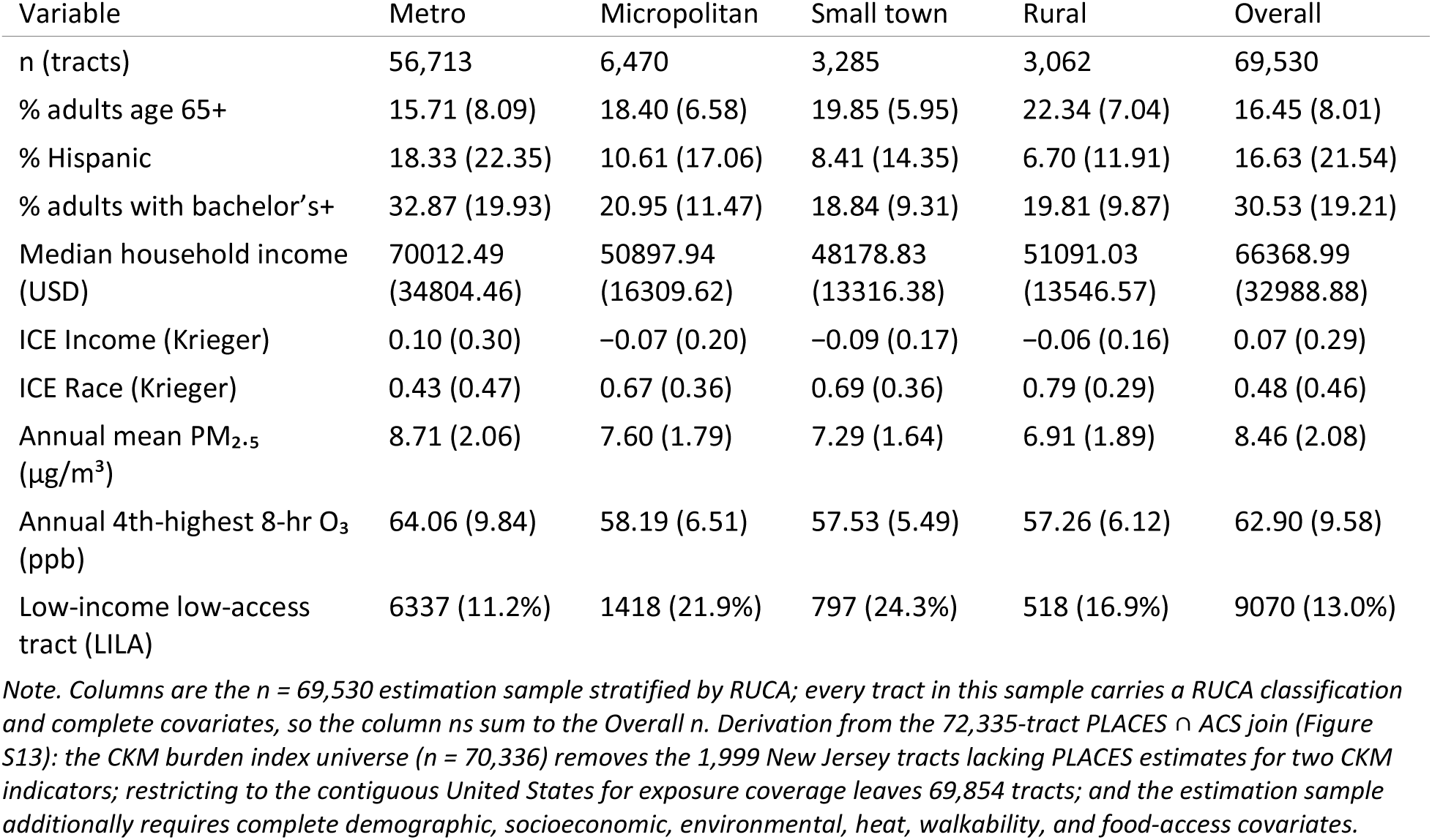
Sample characteristics of the n = 69,530 estimation sample by rural–urban commuting area (RUCA) stratum; see Figure S13 and the note below for the derivation from the 72,335-tract CDC PLACES ∩ ACS attribute join. Continuous variables: mean (SD); binary variables: n (%).

| Variable | Metro | Micropolitan | Small town | Rural | Overall |
|---|---|---|---|---|---|
| n (tracts) | 56,713 | 6,470 | 3,285 | 3,062 | 69,530 |
| % adults age 65+ | 15.71 (8.09) | 18.40 (6.58) | 19.85 (5.95) | 22.34 (7.04) | 16.45 (8.01) |
| % Hispanic | 18.33 (22.35) | 10.61 (17.06) | 8.41 (14.35) | 6.70 (11.91) | 16.63 (21.54) |
| % adults with bachelor’s+ | 32.87 (19.93) | 20.95 (11.47) | 18.84 (9.31) | 19.81 (9.87) | 30.53 (19.21) |
| Median household income (USD) | 70012.49 (34804.46) | 50897.94 (16309.62) | 48178.83 (13316.38) | 51091.03 (13546.57) | 66368.99 (32988.88) |
| ICE Income (Krieger) | 0.10 (0.30) | −0.07 (0.20) | −0.09 (0.17) | −0.06 (0.16) | 0.07 (0.29) |
| ICE Race (Krieger) | 0.43 (0.47) | 0.67 (0.36) | 0.69 (0.36) | 0.79 (0.29) | 0.48 (0.46) |
| Annual mean PM<sub>2.5</sub> (µg/m<sup>3</sup>) | 8.71 (2.06) | 7.60 (1.79) | 7.29 (1.64) | 6.91 (1.89) | 8.46 (2.08) |
| Annual 4th-highest 8-hr O<sub>3</sub> (ppb) | 64.06 (9.84) | 58.19 (6.51) | 57.53 (5.49) | 57.26 (6.12) | 62.90 (9.58) |
| Low-income low-access tract (LILA) | 6337 (11.2%) | 1418 (21.9%) | 797 (24.3%) | 518 (16.9%) | 9070 (13.0%) |

*Note. Columns are the n = 69,530 estimation sample stratified by RUCA; every tract in this sample carries a RUCA classification and complete covariates, so the column ns sum to the Overall n. Derivation from the 72,335-tract PLACES ∩ ACS join (Figure S13): the CKM burden index universe (n = 70,336) removes the 1,999 New Jersey tracts lacking PLACES estimates for two CKM indicators; restricting to the contiguous United States for exposure coverage leaves 69,854 tracts; and the estimation sample additionally requires complete demographic, socioeconomic, environmental, heat, walkability, and food-access covariates.*

**Table 2.** Comparison of tract-level CDC PLACES with individual-level NHANES 2017–2020 for the five CKM conditions: loadings on the first principal component (PC1) of the CKM burden index, and national weighted prevalences. NHANES prevalences are individual-weighted; PLACES prevalences are tract-population-weighted. Difference is NHANES minus PLACES, in percentage points, and is computed from unrounded values, so it may differ in the last decimal place from the difference of the two rounded prevalences shown.

| Condition | PC1 loading, NHANES | PC1 loading, PLACES | Prevalence, NHANES (%) | Prevalence, PLACES (%) | Difference (pct. points) |
|---|---|---|---|---|---|
| Diabetes mellitus | 0.509 | 0.486 | 11.60 | 10.38 | 1.22 |
| Hypertension | 0.576 | 0.494 | 31.85 | 31.55 | 0.30 |
| Obesity | 0.326 | 0.394 | 41.44 | 32.49 | 8.95 |
| Hypercholesterolemia | 0.483 | 0.364 | 33.67 | 31.43 | 2.23 |
| Chronic kidney disease | 0.263 | 0.482 | 3.21 | 2.84 | 0.37 |

We compared PLACES with individual-level NHANES 2017–2020 data (Table 2, Figure 1; Supplemental Results). National prevalences were similar for diabetes, hypertension, hypercholesterolemia, and chronic kidney disease, differing by 0.30 to 2.23 percentage points. Obesity was 8.95 percentage points lower in PLACES, as expected, because BRFSS relies on self-reported height and weight, whereas NHANES measures them. The relationships among the indicators agreed less well. When we compared the ten pairwise correlations in PLACES with those in NHANES, they agreed only weakly (r = 0.37), and the PC1 loadings agreed even less (r = 0.16). This is consistent with how PLACES estimates are produced: the five indicators are much more correlated in PLACES (mean |r| = 0.69) than in NHANES (mean |r| = 0.16), and all ten pairs are more correlated in PLACES (Supplemental Note 2).

**Figure 1.**
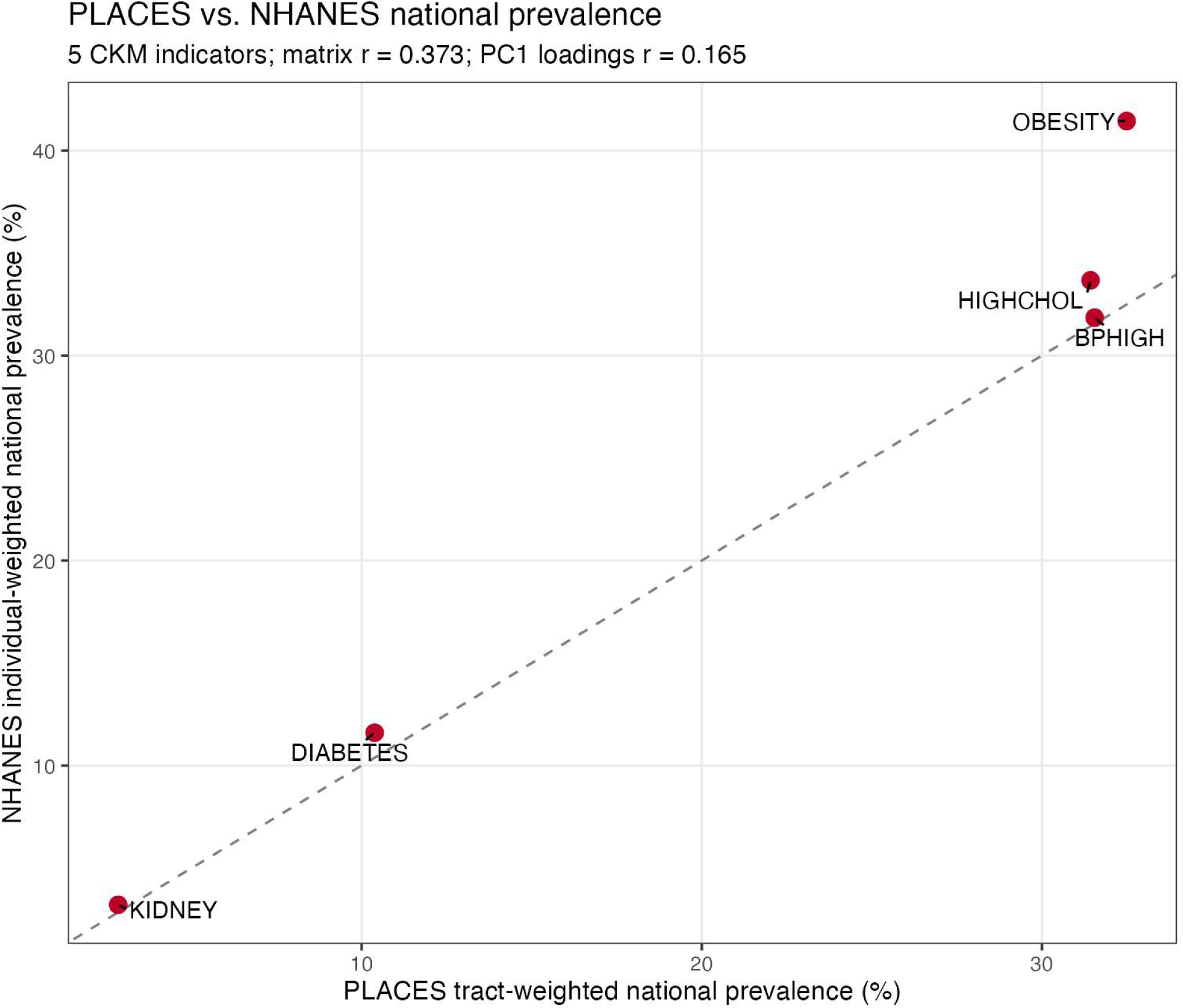
Comparison with NHANES: PLACES vs. NHANES national prevalences for five CKM indicators.

### 3.2 OLS baseline and global Moran’s I diagnostics

The baseline OLS model included only the demographic and socioeconomic predictors (n = 69,530) and explained 72.4% of the variance. Older neighborhoods had higher burden, and both ICE measures had the expected protective (negative) sign. When the behavioral and healthcare-engagement block is added, both ICE coefficients change sign; §3.3 examines this change. Supplemental Results reports the coefficients and the collinearity and normality checks (Figures S11–S12). Figure 2 maps % adults aged ≥65 years, ICE_Income, and ICE_Race.

**Figure 2.**
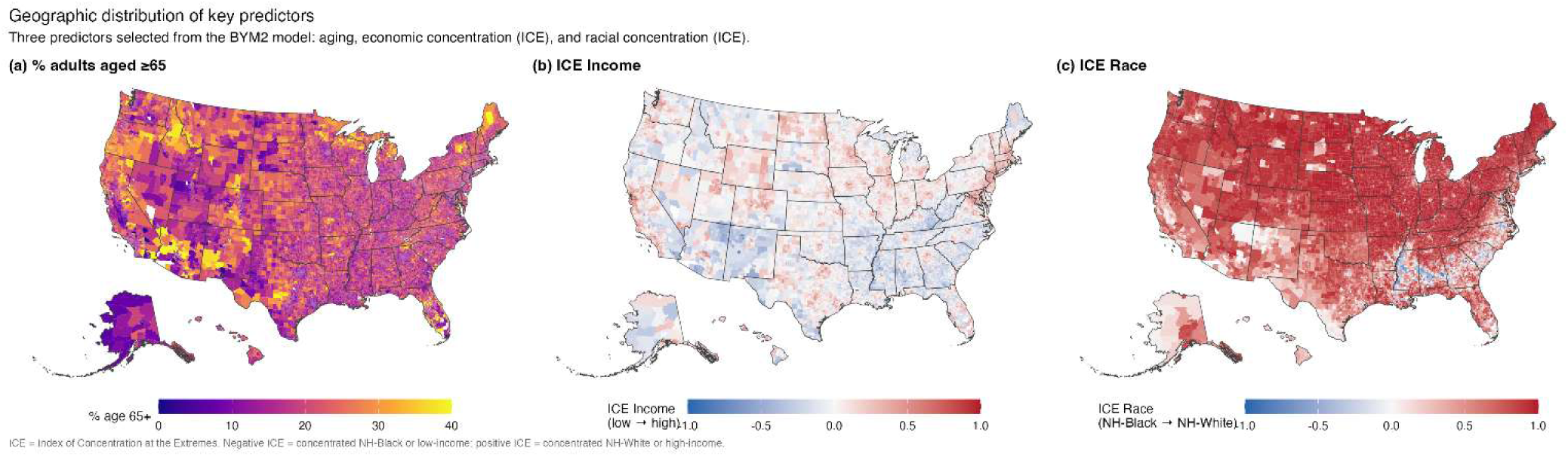
Geographic distribution of three key predictors: percentage of adults aged ≥65 years and the Index of Concentration at the Extremes (ICE) for income and for race.

CKM burden was clustered in space. Global Moran’s I was 0.657 (p < .001) across the 69,854 contiguous-U.S. tracts with a burden index (Figure S13), using row-standardized weights based on each tract’s 8 nearest neighbors. Burden was highest across the U.S. South, including the Mississippi Delta, the Black Belt, Appalachia, and the Rio Grande Valley. It was lowest in the Mountain West, the Pacific Northwest, and the urban core of the Northeast (Figure 3). The OLS residuals were also clustered (Moran’s I = 0.470, p < .001; Figure S1): observed burden was higher than the model predicted in South Texas, the Mississippi Delta, and Appalachia. This remaining clustering is why we used a model with spatial random effects.

**Figure 3.**
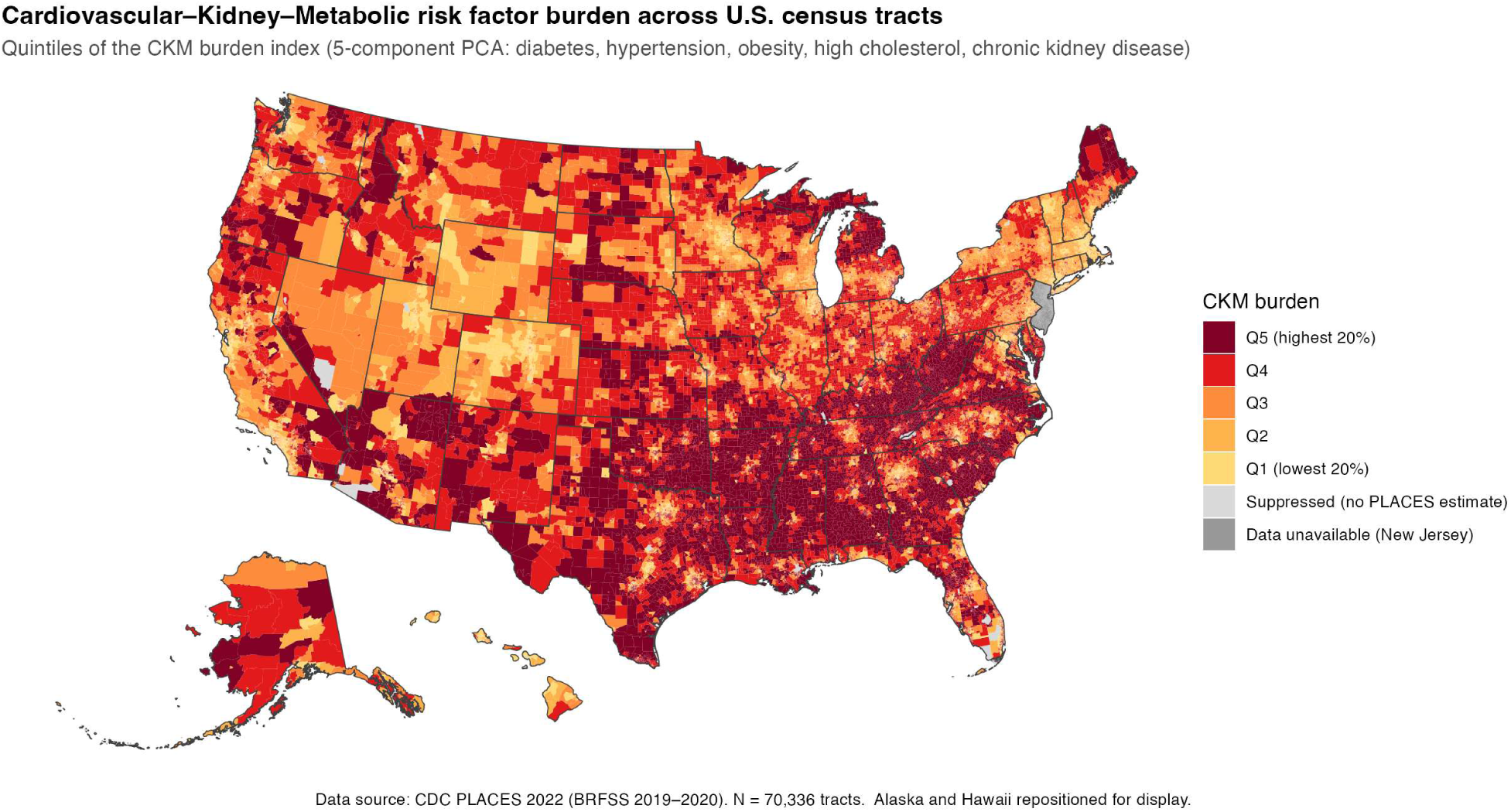
Geography of CKM burden across U.S. census tracts (quintiles of the CKM burden index). New Jersey (gray) lacks PLACES indicator data and is not modeled.

Supplemental Results (Figures S7–S10) reports local clusters (LISA), a correlogram by lag order, and robust Lagrange multiplier tests. Both robust tests, for spatial lag and for spatial error, were significant, suggesting that both types of spatial dependence are present.

### 3.3 BYM2 fit, ESF cross-check, and residual absorption

We fit the BYM2 model on all 69,530 tracts; no further tracts were excluded. Log marginal likelihood was higher for Model 2 (−20,271 vs −73,551; Table S15), but we do not use this difference to choose between the models, because Model 2 includes predictors that come from the same BRFSS data as the outcome (§S2.4; §4.5). Table 3 gives the estimates for both models, Figure 4 compares them, and Table S2 gives the OLS and ESF cross-check estimates for the Model 2 predictor set.

**Figure 4.**
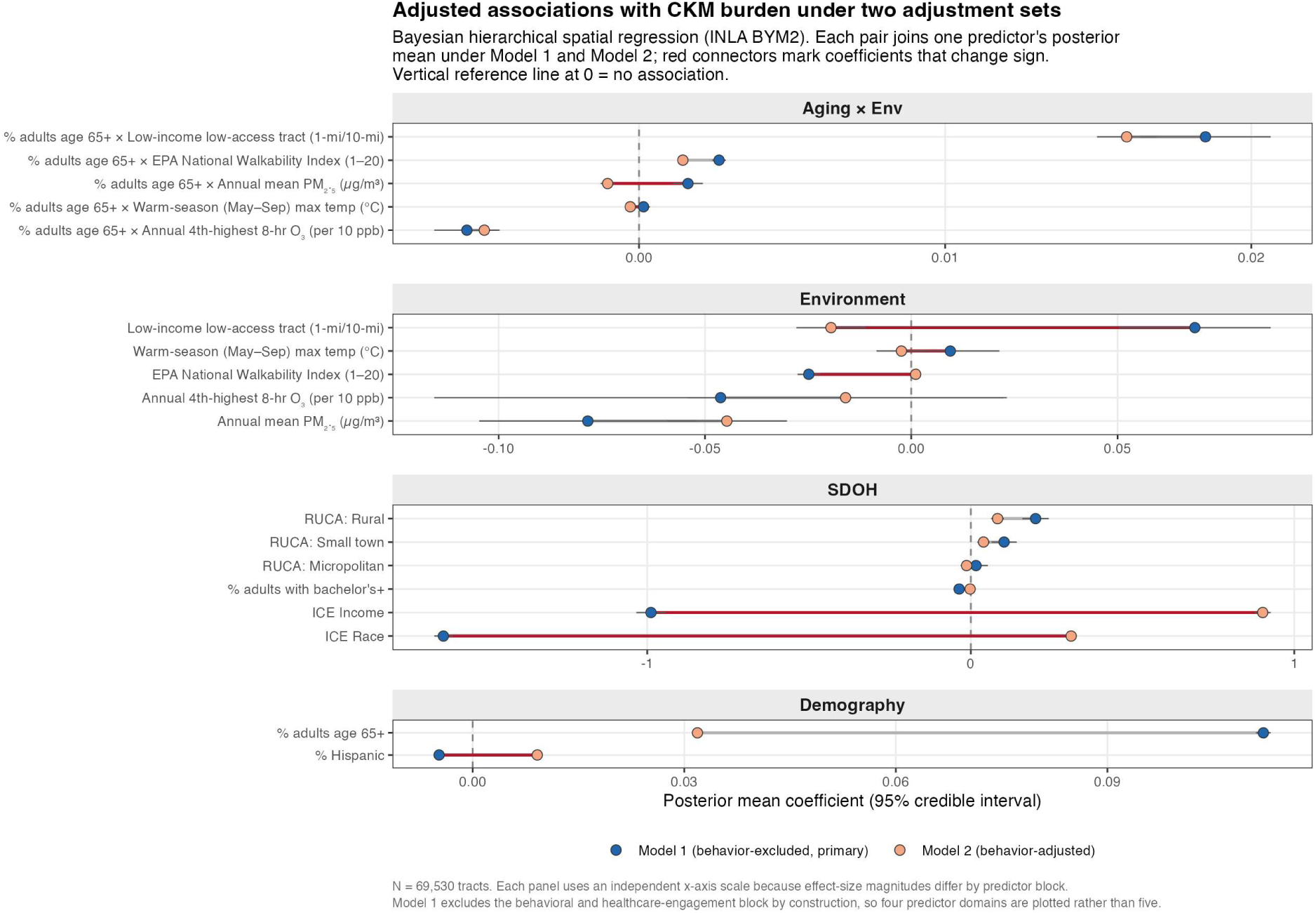
Adjusted associations of neighborhood characteristics with CKM burden under two adjustment sets (BYM2 spatial models). Each row joins one predictor’s posterior mean under Model 1 (behavior-excluded; the primary specification for interpretation, §S2.4) and Model 2 (behavior-adjusted), shown with 95% credible intervals; red connectors mark the nine coefficients that change sign between specifications. Model 1 excludes the behavioral and healthcare-engagement block (CSMOKING, LPA, CHECKUP, ACCESS2) by construction, so four predictor domains are plotted rather than five and its coefficients are behavior-excluded rather than behavior-adjusted associations. Each panel uses an independent x-axis scale because effect-size magnitudes differ by predictor block, and coefficients should therefore not be ranked across panels; for the same reason the attenuation of educational attainment is not resolvable at the SDOH panel scale. Exact posterior summaries for every term are given in Table 3.

**Table 3.**
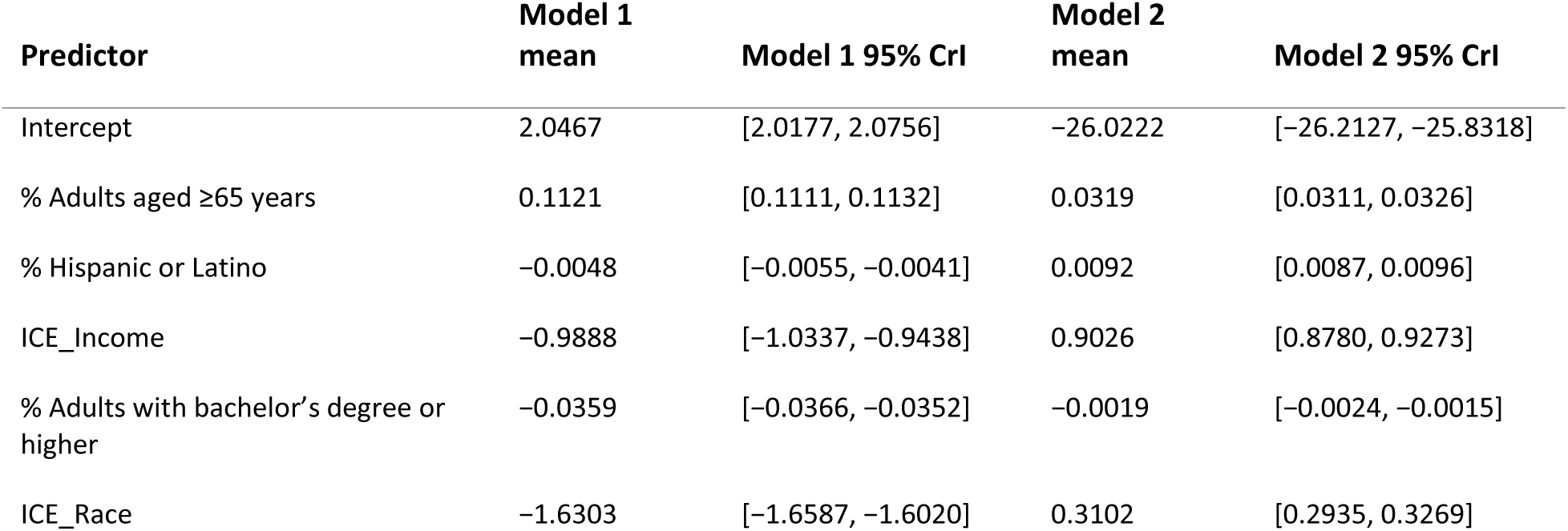
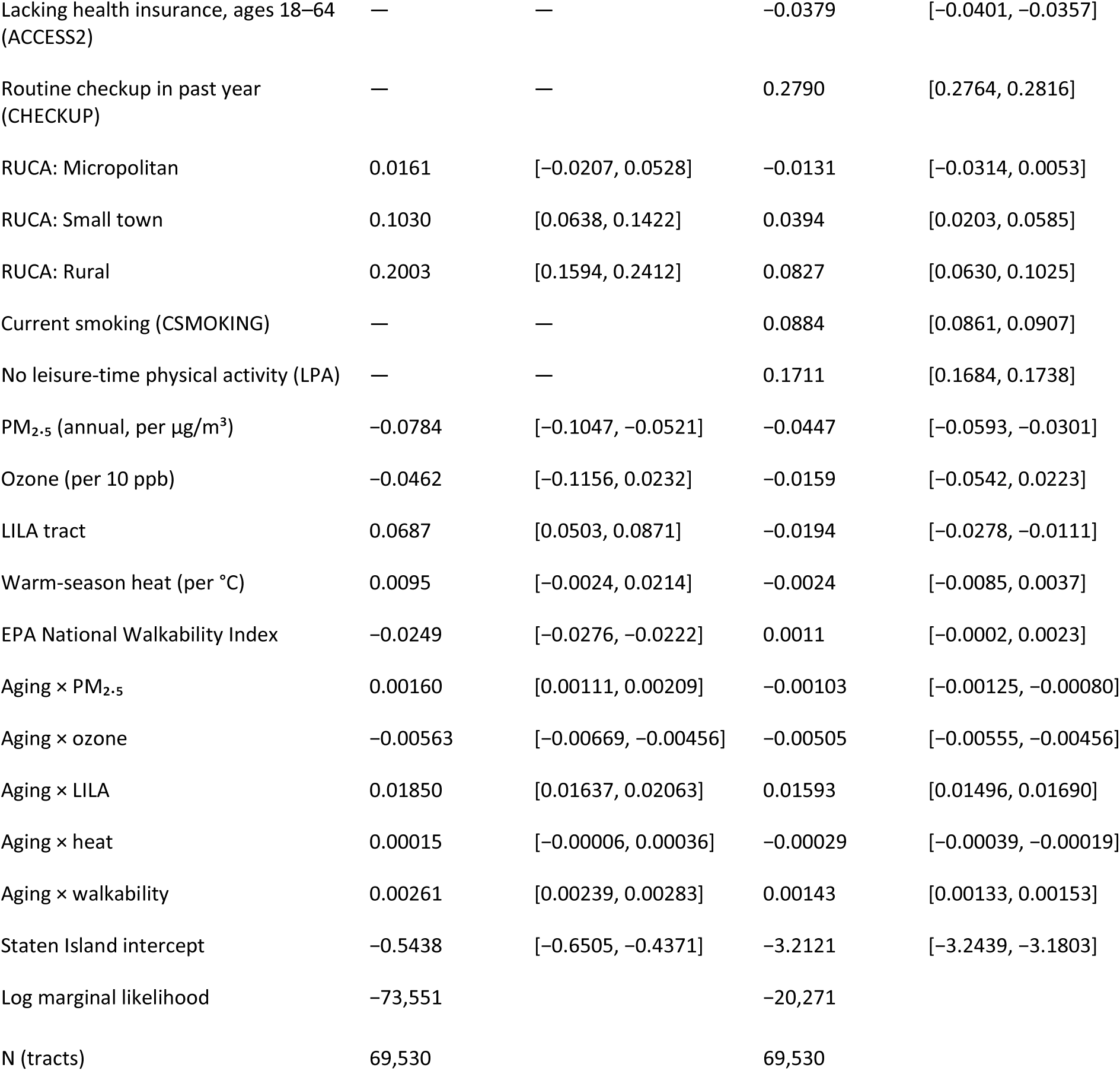
**BYM2 posterior coefficients without (Model 1) and with (Model 2) the behavioral and healthcare-engagement block, fit on the same 69,530 contiguous-U.S. tracts by integrated nested Laplace approximation (INLA).** Model 1 is the primary specification for interpretation (§S2.4): it carries the demographic, socioeconomic, and environmental covariates plus the aging × environment interactions, and excludes the four co-reported behavioral and healthcare-engagement indicators (no leisure-time physical activity, current smoking, routine checkup, and lack of health insurance). Model 2 is the otherwise identical fully adjusted specification (same neighborhood graph and priors) that adds those four indicators; it is retained as the stated conditional contrast rather than as a sensitivity analysis. Entries are posterior means with 95% credible intervals (CrI); continuous predictors entering an interaction are mean-centered and ozone is scaled to units of 10 ppb (§S2.4), and RUCA coefficients are contrasts against Metropolitan tracts. Model 1 is nested within Model 2 (it is Model 2 without the four behavioral and healthcare-engagement coefficients), and the two are presented side by side as readings of the structural and environmental associations under different adjustment sets, rather than as a formal model comparison or as bounds on a single underlying quantity; a dash (—) marks predictors excluded from Model 1. When the behavioral block is removed, both concentration measures and % Hispanic revert to their expected protective (negative) direction, so the positive fully-adjusted coefficients depend on conditioning on the co-reported behavioral block; for ICE_Race and % Hispanic, whose full-covariate OLS coefficients stay negative (Table S2), the spatial random effect also contributes (§3.3). Ordinary least squares (OLS) and eigenvector spatial filter (ESF) estimates for the Model 2 specification are reported in Table S2; the OLS baseline (n = 69,530; §3.2) is reported in Supplemental Results and Table S1. The Staten Island intercept is a component-level offset for the 106 tracts that form a separate part of the neighbor graph (§S2.4): the BYM2 term sums to zero within each part, so this term carries that part’s mean level, and it is not interpreted substantively. It is larger in Model 2 (−3.21, about 1.7 outcome SDs) than in Model 1 (−0.54) because the two models differ by the behavioral block, which changes the fixed-effect fit for these tracts. It contributes +0.008 to the South-minus-non-South difference (Table S11).

| Predictor | Model 1 mean | Model 1 95% CrI | Model 2 mean | Model 2 95% CrI |
|---|---|---|---|---|
| Intercept | 2.0467 | [2.0177, 2.0756] | −26.0222 | [−26.2127, −25.8318] |
| % Adults aged ≥65 years | 0.1121 | [0.1111, 0.1132] | 0.0319 | [0.0311, 0.0326] |
| % Hispanic or Latino | −0.0048 | [−0.0055, −0.0041] | 0.0092 | [0.0087, 0.0096] |
| ICE_Income | −0.9888 | [−1.0337, −0.9438] | 0.9026 | [0.8780, 0.9273] |
| % Adults with bachelor’s degree or higher | −0.0359 | [−0.0366, −0.0352] | −0.0019 | [−0.0024, −0.0015] |
| ICE_Race | −1.6303 | [−1.6587, −1.6020] | 0.3102 | [0.2935, 0.3269] |
| Lacking health insurance, ages 18–64 (ACCESS2) | — | — | −0.0379 | [−0.0401, −0.0357] |
| Routine checkup in past year (CHECKUP) | — | — | 0.2790 | [0.2764, 0.2816] |
| RUCA: Micropolitan | 0.0161 | [−0.0207, 0.0528] | −0.0131 | [−0.0314, 0.0053] |
| RUCA: Small town | 0.1030 | [0.0638, 0.1422] | 0.0394 | [0.0203, 0.0585] |
| RUCA: Rural | 0.2003 | [0.1594, 0.2412] | 0.0827 | [0.0630, 0.1025] |
| Current smoking (CSMOKING) | — | — | 0.0884 | [0.0861, 0.0907] |
| No leisure-time physical activity (LPA) | — | — | 0.1711 | [0.1684, 0.1738] |
| PM<sub>2.5</sub> (annual, per µg/m<sup>3</sup>) | −0.0784 | [−0.1047, −0.0521] | −0.0447 | [−0.0593, −0.0301] |
| Ozone (per 10 ppb) | −0.0462 | [−0.1156, 0.0232] | −0.0159 | [−0.0542, 0.0223] |
| LILA tract | 0.0687 | [0.0503, 0.0871] | −0.0194 | [−0.0278, −0.0111] |
| Warm-season heat (per °C) | 0.0095 | [−0.0024, 0.0214] | −0.0024 | [−0.0085, 0.0037] |
| EPA National Walkability Index | −0.0249 | [−0.0276, −0.0222] | 0.0011 | [−0.0002, 0.0023] |
| Aging × PM<sub>2.5</sub> | 0.00160 | [0.00111, 0.00209] | −0.00103 | [−0.00125, −0.00080] |
| Aging × ozone | −0.00563 | [−0.00669, −0.00456] | −0.00505 | [−0.00555, −0.00456] |
| Aging × LILA | 0.01850 | [0.01637, 0.02063] | 0.01593 | [0.01496, 0.01690] |
| Aging × heat | 0.00015 | [−0.00006, 0.00036] | −0.00029 | [−0.00039, −0.00019] |
| Aging × walkability | 0.00261 | [0.00239, 0.00283] | 0.00143 | [0.00133, 0.00153] |
| Staten Island intercept | −0.5438 | [−0.6505, −0.4371] | −3.2121 | [−3.2439, −3.1803] |
| Log marginal likelihood | −73,551 |  | −20,271 |  |
| N (tracts) | 69,530 |  | 69,530 |  |

#### Demography

Population aging was the strongest demographic predictor. In the XGBoost model, it accounted for 19.3% of the total SHAP contribution, second only to physical inactivity (§S3.4). In the BYM2 models, its association was positive in both models. In Model 1, each 1-percentage-point increase in adults aged ≥65 years was associated with 0.112 units higher burden (95% CrI 0.111–0.113) in non-LILA tracts at average levels of the other exposures. This is a linear average: a natural spline for aging fit better (log marginal likelihood −69,642 vs −73,551), and 3,776 of that 3,909-unit gain came from the aging main effect (Table S13), so the age gradient is not constant. The association was weaker in Model 2 (Table 3; Figure 4). Across the middle 90% of tracts (6% to 29% aged ≥65 years), this slope means a difference of about 2.5 index units, or about 1.3 standard deviations of the outcome. On a per-standard-deviation scale, aging was the largest single effect in Model 1 (+0.90, or 46.5% of an outcome standard deviation; Table S3).

#### Structural socioeconomic conditions

In Model 1, both ICE measures had the expected protective sign (ICE_Income β = −0.99 [−1.03, −0.94]; ICE_Race β = −1.63 [−1.66, −1.60]), as they did in the OLS baseline (§3.2) and the county-level re-analysis (§S3.5). In Model 2, which adds the behavioral and healthcare-engagement block, both became positive (Table 3; Figure 4). Percent Hispanic or Latino changed in the same way, from −0.005 [−0.005, −0.004] to +0.009, and the coefficient for a bachelor’s degree or higher shrank almost to zero (from −0.0359 to −0.0019). For ICE_Income, the sign change also appears in the OLS and ESF models with the Model 2 predictors (Table S2), so it follows from adding the behavioral block. For ICE_Race, OLS and ESF with the same predictors stayed negative (β = −0.80 and −0.07), so the spatial random effect also contributes to its sign change. The data cannot tell us why adding the behavioral block changes these estimates. The behavioral measures come from the same BRFSS data as the outcome and may lie on the pathway between structural disadvantage and CKM burden. Adjusting for them could therefore cause over-adjustment, common-method bias, or collider bias; alternatively, Model 1 may be confounded by factors that the behavioral block captures (§S2.4). Supplemental Results reports the full comparison.

#### Rurality

Burden rose from metropolitan to rural tracts in both models. In Model 1, rural tracts had higher burden than metropolitan tracts (β = +0.200 [+0.159, +0.241]), small-town tracts less so, and micropolitan tracts did not differ from metropolitan tracts (Table 3; Figure 4).

#### Environmental exposures

Figure 5 maps PM₂.₅, and Figures S4–S6 map ozone, warm-season heat, and walkability. The environmental main effects depended on which model was used. PM₂.₅ was negative in both models, but only 0.6% of its variation in Model 1 and 0.1% in Model 2 remained after the spatial term (Table S15), so we do not interpret its sign. Supplemental Results reports ozone, warm-season heat, walkability, and the LILA flag.

**Figure 5.**
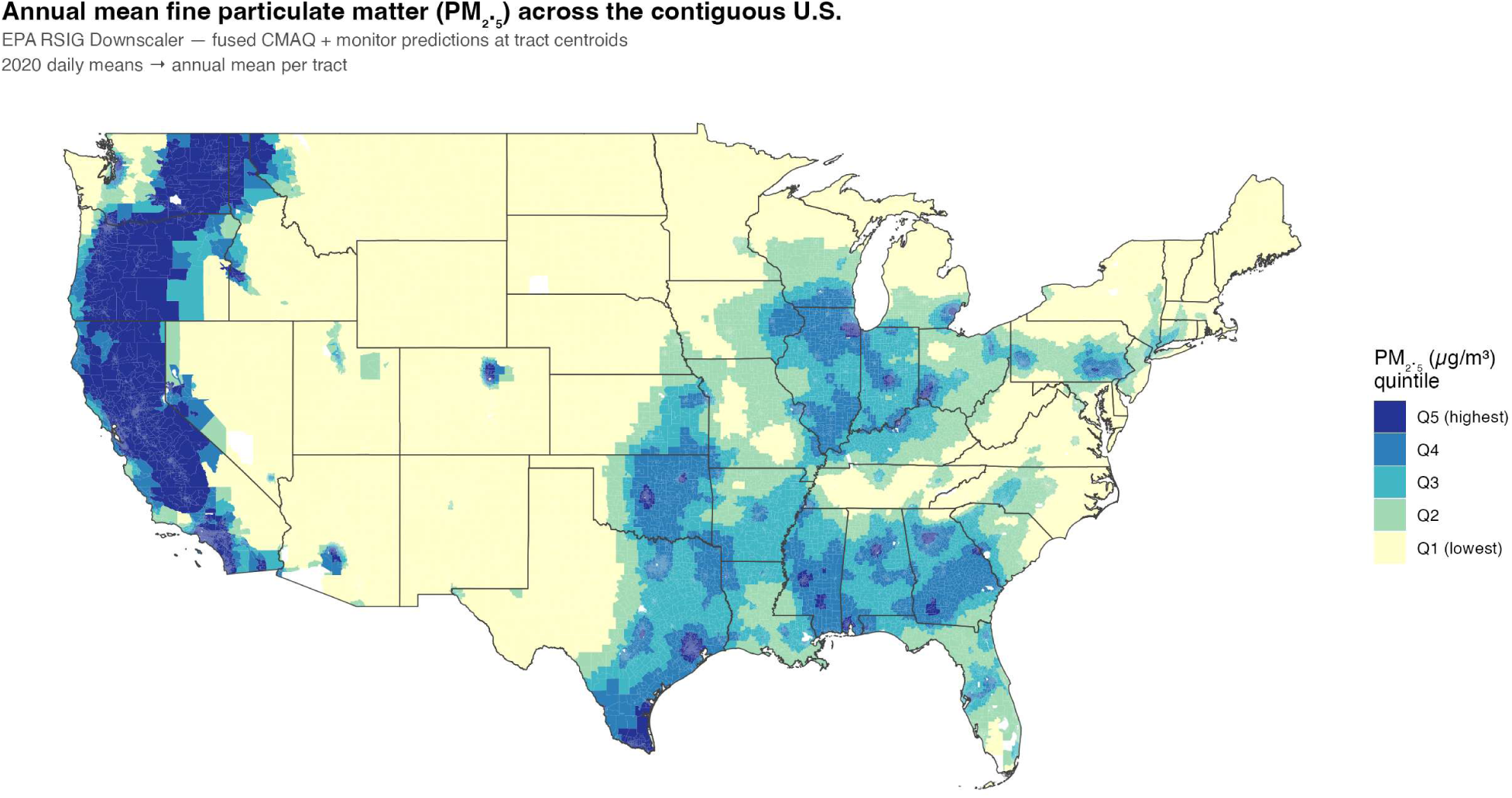
Annual mean PM₂.₅ across the contiguous U.S., year 2020 (EPA RSIG Downscaler).

#### Aging × environment interactions

With aging entered linearly, the aging × LILA interaction was positive (Model 1 β = +0.019 [+0.016, +0.021]; Model 2 β = +0.016 [+0.015, +0.017]). This linear form implies that the LILA difference keeps rising with age composition and becomes negative in the youngest tracts. A natural spline for aging (df = 4) fit better (log marginal likelihood −69,642 vs −73,551) and showed a different shape: LILA tracts had 0.10 to 0.14 units higher burden from the 25th percentile of aging upward, with no credible difference in the youngest decile (Table S13). However, 97% of this gain came from the aging main effect: a spline main effect with a linear interaction recovered 3,776 of the 3,909, and freeing the interaction added 132. The shape in the youngest tracts therefore depends on the functional form. With food-store counts per resident in place of the LILA flag, the higher burden above the 25th percentile of aging did not appear, and adding the store counts left the LILA contrasts nearly unchanged (§S3.10; Table S14). The other four interactions were small (Table 3; Figure 4). Aging × ozone was negative and aging × walkability was positive in both models. Aging × PM₂.₅ (positive in Model 1) and aging × heat (not different from zero in Model 1) both became negative in Model 2.

Supplemental Results reports the ESF cross-check, the comparison of fit across modeling frameworks (Table S4), the focal-versus-neighborhood (Wx) analysis (Tables S5–S6), and the Southern excess that remains after adjustment, with its dependence on the model (Figure 6; Table S11).

**Figure 6.**
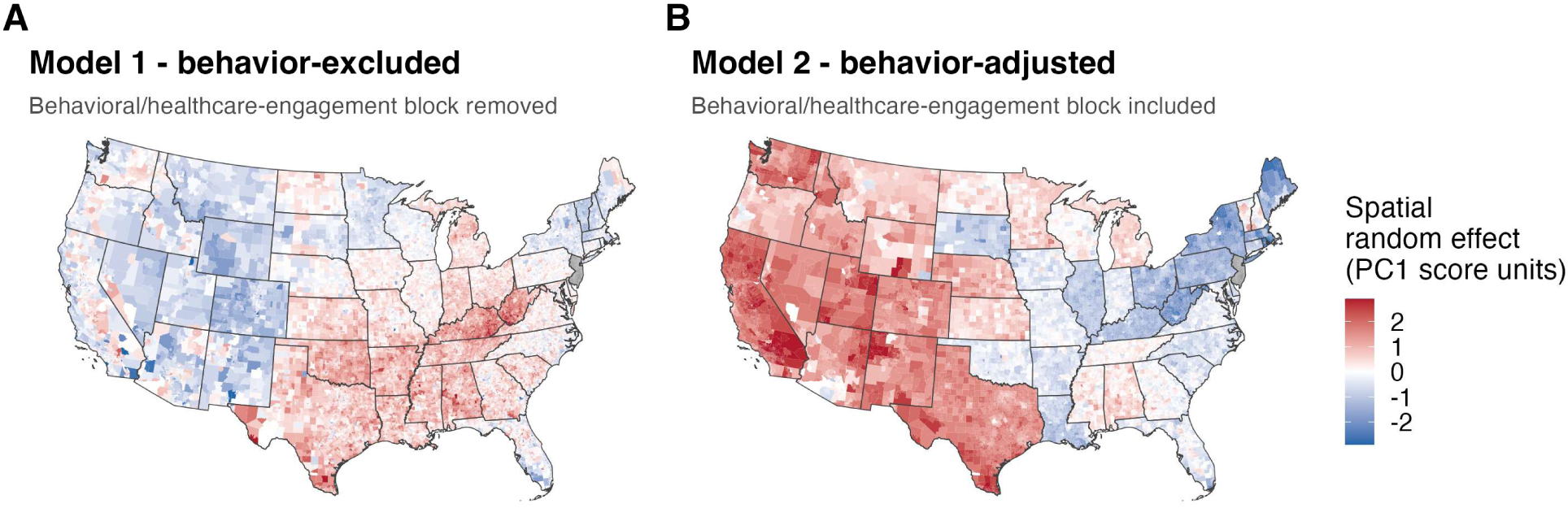
Residual spatial structure in CKM burden under two adjustment sets. Both panels map the posterior mean BYM2 spatial random effect (PC1 component-score units) across the 69,530 contiguous-U.S. tracts on the shared k = 8 neighbor graph, on a common color scale; red denotes burden in excess of what the measured predictors explain. (A) Model 1, behavior-excluded, the primary specification: demographic, socioeconomic, environmental, and rurality predictors, without the behavioral and healthcare-engagement block. (B) Model 2, behavior-adjusted: identical neighborhood graph, priors, and covariates with that block (physical inactivity, current smoking, routine checkup, and health-insurance access) added.The Southern excess across the Mississippi Delta, the Black Belt, and Appalachia is present in Model 1 and absent in Model 2, whereas Texas remains positive in both (state mean random effect +0.58 in Model 1 and +1.52 in Model 2); because the behavioral block consists of PLACES small-area estimates that share a BRFSS survey basis with the CKM outcome, this contrast may reflect shared method variance and diagnostic ascertainment, so we do not treat it as an explanation of the regional gradient (§3.3, §4.1, §4.5). New Jersey (gray) lacks PLACES indicator data and is not modeled.

### 3.4 Sensitivity analyses

Supplemental Results reports seven sensitivity analyses in full: county-level re-aggregation (Table S8, Panel A), alternative spatial weights (Table S8, Panel B), aging × environment interactions refit within rurality strata (Table S8, Panel C), an index without chronic kidney disease, refit for Model 2 (Table S9), precision weighting using the CDC PLACES confidence intervals (Table S10), an extended index that adds coronary heart disease and stroke, refit for Model 1 (§S3.9), and food-store counts in place of the LILA flag, refit for Model 1 (§S3.10; Table S14). Four of these analyses re-estimate the main effects: county re-aggregation, the index without chronic kidney disease, precision weighting, and the extended index. In all four, the main effects of population aging and the two ICE measures kept their direction. The aging × environment interactions were less stable. Aging × heat changed the most: it was not different from zero in the primary tract-level model, but it was negative (nominally significant) at the county scale (§S3.5) and in every rurality stratum (§S3.8), and positive in the precision-weighted and extended-index refits of Model 1 (§S3.7, §S3.9). The county analysis tests whether results depend on the spatial scale of analysis, which is one part of the Modifiable Areal Unit Problem.

## 4. Discussion

### 4.1 Principal findings

The tract-level index of AHA Stage 1–2 CKM burden was clustered in space (§3.2) and agreed only modestly with NHANES individual-level data. Between the two sources, the correlations among the five indicators agreed at r = 0.37, and the PC1 loadings at r = 0.16. Chronic kidney disease accounted for most of the disagreement: its PC1 loading was 0.48 in PLACES but 0.26 in NHANES. This gap may partly reflect a difference in case definition, because NHANES asks about “weak or failing kidneys” (§S2.2), but it is also consistent with the model-based smoothing behind PLACES tract estimates. We highlight three main findings.

First, the behavioral and healthcare-engagement measures ranked highest in the descriptive XGBoost analysis, with 52.6% of total SHAP importance (§S3.4; Figure 7). Physical inactivity ranked first, and smoking and routine checkup were among the top four features; population aging ranked second and was the leading demographic feature. These measures come from the same BRFSS/PLACES data as the outcome. Their high ranking therefore at least partly reflects common-method variance and diagnostic ascertainment (§4.5), not an ordering of causes, and we read them as descriptive correlates. In the same model, the socioeconomic measures (ICE_Income, ICE_Race, education, and rurality) accounted for 11.5% of total importance. On a per-standard-deviation scale, Model 2, which uses the same predictors as XGBoost, ranked the same two leading blocks first, but the order of the environmental and socioeconomic blocks differed (Table S3).

**Figure 7.**
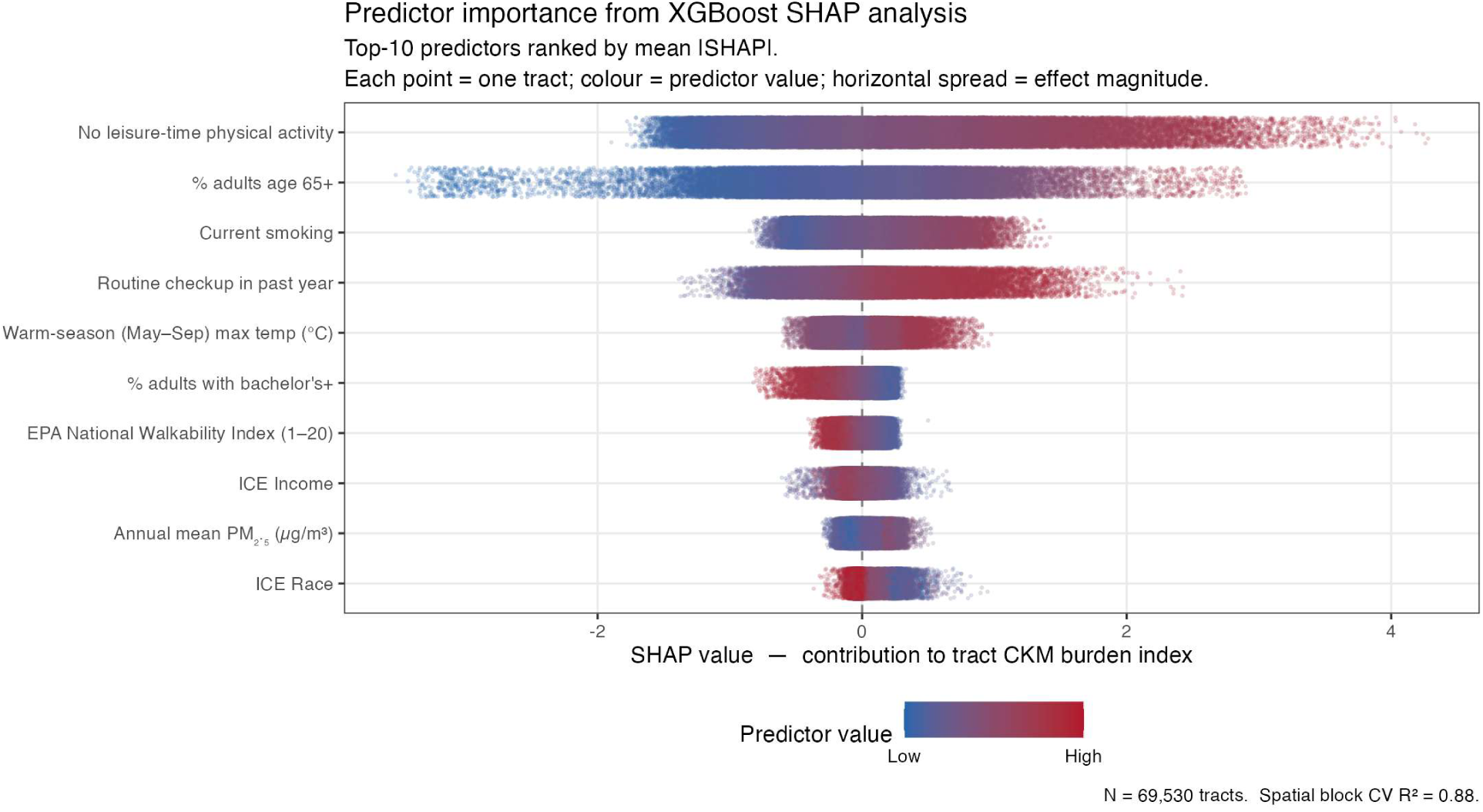
Relative importance of predictors of CKM burden in the machine-learning model (top 10), summarized by SHAP (SHapley Additive exPlanations) values; shown as a descriptive complement to the spatial-regression results.

Second, environmental associations were modest, and evidence that they vary with age composition was limited. In the primary model, LILA tracts had 0.10 to 0.14 units higher burden from the 25th percentile of aging upward, with no credible difference in the youngest tracts; the shape in the youngest tracts depends on functional form (§3.3). This LILA pattern was also specific to the USDA flag, which combines low income with distance to a supermarket; food-store counts per resident did not reproduce it (§S3.10). The aging × PM₂.₅ interaction was positive in the primary model and matched the metropolitan stratum in sign (§4.3). The aging × ozone interaction was negative in the pooled models but reversed sign between metropolitan and small-town/rural tracts (Table S8; §4.3). The aging × walkability interaction was small and positive in all three rurality strata, although it reversed sign at the county scale (§S3.5). The aging × heat interaction was the least robust. It was negative in every rurality stratum, in Model 2, and at the county scale, but not different from zero in the primary pooled model and positive in the precision-weighted, spatial+, and extended-index refits of Model 1 (§S3.7, §S3.9; Table S12). Its direction is therefore unresolved. The SHAP dependence plots support these estimates only in part (§S3.4; Figure 8).

**Figure 8.**
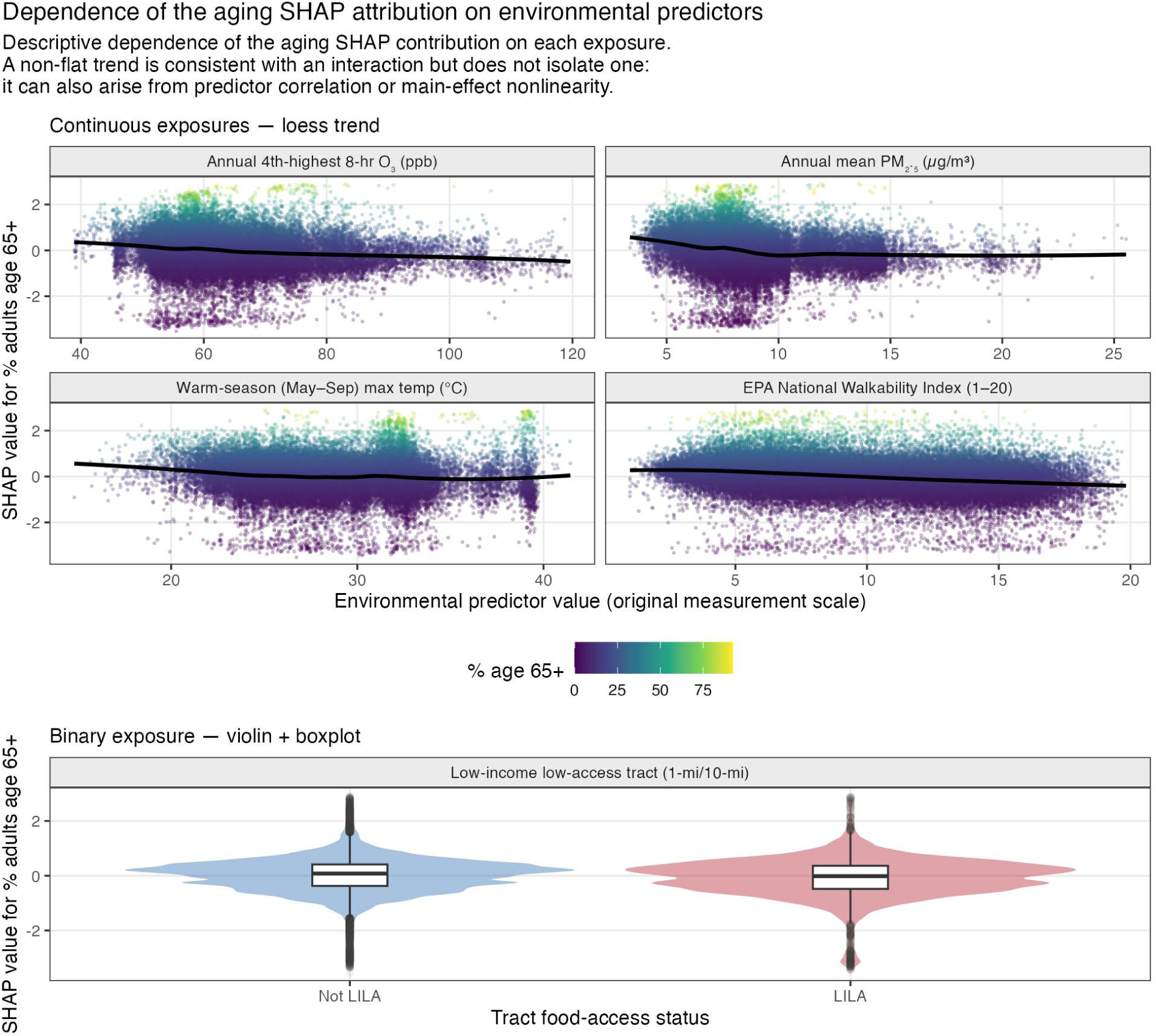
Dependence of the aging SHAP attribution on environmental predictors: machine-learning (SHAP) dependence plots. The vertical axis in both sections is the SHAP value for percent adults aged 65 and older. The upper section shows the four continuous exposures (annual 4th-highest 8-hour ozone, annual mean PM₂.₅, warm-season maximum temperature for May–September, and the EPA National Walkability Index) as SHAP scatter with a loess trend, each point colored by tract percent adults aged 65 and older; a non-flat trend is consistent with an age-by-exposure interaction but does not isolate one, since it can also arise from predictor correlation or main-effect nonlinearity. The lower section shows the one binary exposure, low-income low-access (LILA) food status (1-mile/10-mile), as a violin plot with an inset boxplot; there the same pattern appears as a shift in the SHAP distribution between the non-LILA and LILA levels rather than by a trend.

Third, the Southern excess in CKM burden depended on the adjustment set. In Model 1, which excludes the behavioral block, the Southern regions named in §3.2 remained the areas of highest residual burden in the BYM2 spatial random effect (Figure 6). In Model 2, which includes the block, the excess approached zero (§S3.3). The block comes from the same BRFSS survey as the outcome, and its care-engagement measures point in the direction expected from diagnostic ascertainment (routine checkup positive, lack of insurance negative; Table 3; §4.5). We therefore consider Model 1 the more interpretable map of residual geography. The loss of the excess in Model 2 may reflect shared measurement and ascertainment, real regional differences in these behaviors, or both, and the data cannot separate them.

### 4.2 Comparison with prior literature

Our finding that structural socioeconomic conditions are associated with tract-level CKM burden, after adjustment for environmental exposures, is consistent with the neighborhood-effects literature^4–6,20^. In Model 1, which excludes the behavioral block, both ICE measures were protective, in the same direction as that literature (§3.3; Table 3). In Model 2, both were positive but small: per standard deviation, +0.26 for ICE_Income and +0.14 for ICE_Race (Table S3). Because the ICE indices range from −1 to +1, their raw coefficients cannot be compared with effect sizes from other studies, so we compare our results with that literature in direction and ordering, not magnitude. Supplemental Discussion gives the rescaling and the same adjustment sensitivity for percent Hispanic or Latino.

The closest prior study is the nationwide analysis by Krishnan et al. (2026), which compared seven composite neighborhood indices with median household income as predictors of coronary heart disease, stroke, and chronic kidney disease and found similar associations. Our study differs in four ways. First, their primary outcomes, coronary heart disease and stroke, are Stage 4 clinical disease, whereas our index measures the earlier Stage 1–2 CKM burden; they analyzed obesity, hypertension, and diabetes only as separate secondary outcomes. Second, we model a single index aligned with CKM Stages 1–2 and compared with NHANES, rather than comparing social indices with income. Third, they adjusted for tract median age, whereas we used the share of adults aged ≥65 years, the group in which CKM conditions are most common. We also test this share as an effect modifier, and because our outcome is not age-standardized, our aging terms reflect both the age makeup of tracts and age-related risk. Fourth, our BYM2 model accounts for spatial autocorrelation, whereas their non-spatial regressions treat tracts as independent. When we refit our primary model on an extended index that adds their Stage 4 endpoints, every coefficient kept its sign and the Southern excess remained (§S3.9). Our study therefore addresses two questions theirs did not: where Stage 1–2 burden concentrates once spatial dependence is modeled, and whether age composition modifies its environmental associations.

We know of no direct precedent at the U.S. tract scale for the LILA difference, which was present across most of the age distribution but absent in the youngest tracts (§3.3). Older adults’ greater reliance on food sources near home^21,22^ could contribute to this pattern, although the difference did not increase further with age composition above the 25th percentile.

### 4.3 Spatial heterogeneity: scale-dependent interactions, residual geography, and neighborhood effects

The main effects of the ambient exposures (PM₂.₅, ozone, and warm-season heat) are difficult to estimate in this model. These exposures vary mainly across regions, and the BYM2 spatial random effect absorbs broad regional patterns in any smoothly varying predictor. A smooth surface over tract locations (thin-plate spline) explains 92% of the variation in warm-season heat, 91% in ozone, and 88% in PM₂.₅, but only 14% in % adults aged ≥65 years (Table S12). The ambient main effects are therefore estimated mostly from differences within regions, and we do not interpret them as exposure–response relationships. The aging × environment interactions are less affected, because they rely on tract-to-tract differences in age composition within regions of similar exposure. Even so, not all of them held up. When we removed the spatial trend from each covariate (spatial+)^23^, the LILA and walkability interactions remained credible, the ozone and PM₂.₅ interactions lost credibility, and the heat interaction, which was not credible in the primary model, became credible and positive (Table S12). Supplemental Discussion reports the spatial+ and state fixed-effect refits, the rural–urban differences in the ozone and PM₂.₅ interactions, and the neighborhood-effect (Wx) estimates.

The Southern excess that remained in Model 1 raises the question of what this residual geography represents. Unmeasured contextual factors may contribute, such as historical segregation and disinvestment, including the legacy of Home Owners’ Loan Corporation (HOLC) redlining^24^, and regional differences in healthcare access and capacity, such as state Medicaid expansion status in 2019–2020^25^. Our ICE_Race, RUCA, and education measures do not fully capture these factors.

### 4.4 Clinical and public-health implications

CKM burden is geographically patterned at the tract level. For clinicians and health systems, area-level information of this kind may complement, but not replace, the individual risk factors assessed in the clinic. The AHA PREVENT equations already accept an optional ZIP code–level Social Deprivation Index, which improved discrimination only modestly^26^. Whether tract-level CKM burden improves individual risk prediction beyond that term has not been tested.

Physical inactivity and smoking are also more common in the neighborhoods with the greatest burden (§S3.3; highest vs lowest burden quintile: physical inactivity 34.0% vs 17.2%, current smoking 24.0% vs 11.9%). Both were associated with higher burden in Model 2, the only model that includes them (Table 3), and both are established modifiable CKM risk factors^1^. That external evidence, rather than their ranking in our analysis, supports non-pharmacological approaches such as community-based physical-activity and tobacco-cessation programs. The care-engagement measures cannot be read the same way. Higher routine-checkup prevalence was associated with higher measured burden (+0.2790), and lack of health insurance with lower burden (−0.0379; Table 3), a pattern consistent with diagnostic ascertainment, so we do not present them as intervention targets (§4.5).

LILA tracts had modestly higher burden across most of the age distribution (0.10 to 0.14 units, about 5% to 7% of an outcome standard deviation; §3.3). This may help guide area-level needs assessment, but it does not show that pairing screening with food-access interventions would improve individual outcomes. Similarly, the Southern excess in Model 1 supports using geography to plan services, but it does not make geography a cause.

### 4.5 Limitations

Four limitations matter most for interpretation. First, the design is ecological. It compares tracts, not individuals, so our coefficients should not be read as individual-level effects. Second, the outcome and several predictors are partly circular. The CDC PLACES model predicts tract prevalence from age, sex, race and ethnicity, education, and county poverty, which overlap with predictors in our models. Part of the demographic and socioeconomic associations may therefore come from the PLACES model itself, and the index should be read as a proxy for burden, not a measure of clinical CKM co-morbidity. Our credible intervals also treat the PLACES estimates as fixed, so they do not include the uncertainty of the PLACES model (§S3.7).

Third, each model is open to a different bias. Model 1 omits the behavioral block and may be confounded by it. Model 2 adjusts for the block, which shares a data source with the outcome and may lie on the causal pathway, so it may be over-adjusted. Fourth, the routine-checkup and health-insurance estimates may reflect ascertainment and reverse causation. PLACES measures diagnosed conditions, so tracts with better access to care can show higher measured burden at the same level of disease. The cross-sectional design also cannot rule out people moving because of their health. Supplemental Note 2 describes these limitations in full, along with sensitivity to spatial scale, exposure measurement error, multiple comparisons, the crude (not age-standardized) outcome, and the exclusion of New Jersey.

### 4.6 Future directions

Future studies should test whether these patterns hold in later CDC PLACES releases, use difference-in-differences designs to test whether changes in food access precede changes in CKM burden, and use individual-level data, such as NHANES and geocoded cohorts, to separate confounding from mediation (Supplemental Discussion).

### 4.7 Conclusions

At the U.S. census-tract scale, CKM burden is spatially clustered. In the analysis, the behavioral and healthcare-engagement measures ranked highest, followed by population aging, with smaller contributions from environmental exposures and structural socioeconomic conditions. This ranking is not causal since the behavioral measures come from the same BRFSS data as the outcome. LILA tracts had modestly higher burden across most of the age distribution, but evidence that age composition changes environmental associations was limited. A Southern excess in burden remained after adjustment for demographic, socioeconomic, and environmental factors and approached zero only when the shared-source behavioral measures were added, so its causes remain unknown. These estimates support area-level surveillance and needs assessment.

## Acknowledgments

None.

## Data and code availability

All data used in this study are publicly available from CDC PLACES, the American Community Survey, NHANES, the U.S. Environmental Protection Agency, the U.S. Department of Agriculture, the National Neighborhood Data Archive, the PRISM Climate Group, and U.S. Census Bureau TIGER/Line files; Table S1a lists the release of each. The derived output tables behind every reported estimate are available at https://doi.org/10.5281/zenodo.22234206, and the analysis code and documentation are available at https://github.com/batobara/CKM-US-tract-analysis. The code also includes four exploratory steps that are not used in this article: a refit of the aging × environment interactions with the CDC Social Vulnerability Index in place of the ICE measures, a check of missing-value coding in the Food Access Research Atlas low-access population counts, a county-level outcome comparison that needs an outcome file not included in the sources above, and a breakdown of the low-income low-access flag into its components.

## Non-standard Abbreviations and Acronyms

BRFSS: Behavioral Risk Factor Surveillance System
BYM2: Besag-York-Mollié 2 spatial model
CKM: cardiovascular-kidney-metabolic
CrI: credible interval
ESF: eigenvector spatial filtering
HOLC: Home Owners’ Loan Corporation
ICE: Index of Concentration at the Extremes
INLA: integrated nested Laplace approximation
LILA: low-income, low-access tract flag (USDA Food Access Research Atlas)
LISA: local indicators of spatial association
NHANES: National Health and Nutrition Examination Survey
OLS: ordinary least squares
PC1: first principal component
PLACES: Population Level Analysis and Community Estimates (CDC)
RUCA: Rural-Urban Commuting Area
SDOH: social determinants of health
SHAP: Shapley additive explanations
Wx: spatially lagged (neighborhood) predictor values
XGBoost: extreme gradient boosting

## References

1. Ndumele CE, Rangaswami J, Chow SL, Neeland IJ, Tuttle KR, Khan SS, Coresh J, Mathew RO, Baker-Smith CM, Carnethon MR, et al. Cardiovascular-Kidney-Metabolic Health: A Presidential Advisory From the American Heart Association. Circulation. 2023;148:1606– 1635. doi: 10.1161/CIR.0000000000001184

2. Martin SS, Aday AW, Almarzooq ZI, Anderson CAM, Arora P, Avery CL, Baker-Smith CM, Barone Gibbs B, Beaton AZ, Boehme AK, et al. 2024 Heart Disease and Stroke Statistics: A Report of US and Global Data From the American Heart Association. Circulation. 2024;149:e347–e913. doi: 10.1161/CIR.0000000000001209

3. U.S. Census Bureau. Census Bureau Releases New 2020 Census Data on Age, Sex, Race, Hispanic Origin, Households and Housing. https://www.census.gov/newsroom/press-releases/2023/2020-census-demographic-profile-and-dhc.html. 2023. Accessed May 15, 2026.

4. Krieger N, Kim R, Feldman J, Waterman PD. Using the Index of Concentration at the Extremes at multiple geographical levels to monitor health inequities in an era of growing spatial social polarization: Massachusetts, USA (2010-14). Int J Epidemiol. 2018;47:788–819. doi: 10.1093/ije/dyy004

5. Kershaw KN, Osypuk TL, Do DP, De Chavez PJ, Diez Roux AV. Neighborhood-level racial/ethnic residential segregation and incident cardiovascular disease: the multi-ethnic study of atherosclerosis. Circulation. 2015;131:141–148. doi: 10.1161/CIRCULATIONAHA.114.011345

6. Diez Roux AV, Mair C. Neighborhoods and health. Ann N Y Acad Sci. 2010;1186:125–145. doi: 10.1111/j.1749-6632.2009.05333.x

7. Pope CA III, Burnett RT, Thun MJ, Calle EE, Krewski D, Ito K, Thurston GD. Lung cancer, cardiopulmonary mortality, and long-term exposure to fine particulate air pollution. JAMA. 2002;287:1132–1141. doi: 10.1001/jama.287.9.1132

8. Brook RD, Rajagopalan S, Pope CA III, Brook JR, Bhatnagar A, Diez-Roux AV, Holguin F, Hong Y, Luepker RV, Mittleman MA, et al. Particulate matter air pollution and cardiovascular disease: An update to the scientific statement from the American Heart Association. Circulation. 2010;121:2331–2378. doi: 10.1161/CIR.0b013e3181dbece1

9. Bowe B, Xie Y, Li T, Yan Y, Xian H, Al-Aly Z. Particulate Matter Air Pollution and the Risk of Incident CKD and Progression to ESRD. J Am Soc Nephrol. 2018;29:218–230. doi: 10.1681/ASN.2017030253

10. Bowe B, Xie Y, Li T, Yan Y, Xian H, Al-Aly Z. The 2016 global and national burden of diabetes mellitus attributable to PM2.5 air pollution. Lancet Planet Health. 2018;2:e301–e312. doi: 10.1016/S2542-5196(18)30140-2

11. Jiang Z, Zhou Y, Bai R, Lin Y, Shi Y, Zheng X, Yu Y, Cai J, Qin G. Long-term exposure to multiple air pollution and trajectory risk of cardio-renal-metabolic multimorbidity. Atmospheric Environment. 2026;385:122340. doi: 10.1016/j.atmosenv.2026.122340

12. Krishnan V, Huang X, McGowan C, Shah NS, Qureshi F, Minkovitz CS, Kershaw KN, Freedman AA, Miller GE, Lloyd-Jones DM, et al. Neighborhood Indices, Income, and Cardiovascular-Kidney-Metabolic Syndrome at the Census Tract Level. JAMA Netw Open. 2026;9:e266019. doi: 10.1001/jamanetworkopen.2026.6019

13. Centers for Disease Control and Prevention. PLACES: Local Data for Better Health. https://www.cdc.gov/places/. 2022. Accessed May 15, 2026.

14. Stierman B, Afful J, Carroll MD, Chen T-C, Davy O, Fink S, Fryar CD, Gu Q, Hales CM, Hughes JP, et al. National Health and Nutrition Examination Survey 2017–March 2020 Prepandemic Data Files—Development of Files and Prevalence Estimates for Selected Health Outcomes. National Health Statistics Reports No. 158. Hyattsville, MD: National Center for Health Statistics; 2021. doi: 10.15620/cdc:106273

15. U.S. Department of Agriculture, Economic Research Service. Rural-Urban Commuting Area Codes. https://www.ers.usda.gov/data-products/rural-urban-commuting-area-codes. Accessed May 15, 2026.

16. Daly C, Halbleib M, Smith JI, Gibson WP, Doggett MK, Taylor GH, Curtis J, Pasteris PP. Physiographically sensitive mapping of climatological temperature and precipitation across the conterminous United States. Int J Climatol. 2008;28:2031–2064. doi: 10.1002/joc.1688

17. U.S. Environmental Protection Agency. Smart Location Mapping. https://www.epa.gov/smartgrowth/smart-location-mapping. 2021. Accessed October 28, 2026.

18. Melendez R, Clary WM II, Clarke PJ, Noppert GA, Gypin L. National Neighborhood Data Archive (NaNDA): Grocery and Food Stores by Census Tract and ZCTA, United States, 1990-2022. Version 2.1. Ann Arbor, MI: Inter-university Consortium for Political and Social Research; 2026. doi: 10.3886/ICPSR209313.V2

19. U.S. Census Bureau. American Community Survey (ACS). https://www.census.gov/programs-surveys/acs. 2020. Accessed October 28, 2026.

20. Krieger N, Waterman PD, Spasojevic J, Li W, Maduro G, Van Wye G. Public Health Monitoring of Privilege and Deprivation With the Index of Concentration at the Extremes. Am J Public Health. 2016;106:256–263. doi: 10.2105/AJPH.2015.302955

21. Beaulac J, Kristjansson E, Cummins S. A systematic review of food deserts, 1966-2007. Prev Chronic Dis. 2009;6:A105.

22. Walker RE, Keane CR, Burke JG. Disparities and access to healthy food in the United States: A review of food deserts literature. Health & Place. 2010;16:876–884. doi: 10.1016/j.healthplace.2010.04.013

23. Dupont E, Wood SN, Augustin NH. Spatial+: A Novel Approach to Spatial Confounding. Biometrics. 2022;78:1279–1290. doi: 10.1111/biom.13656

24. Aaronson D, Hartley D, Mazumder B. The Effects of the 1930s HOLC “Redlining” Maps. American Economic Journal: Economic Policy. 2021;13:355–392. doi: 10.1257/pol.20190414

25. Khatana SAM, Bhatla A, Nathan AS, Giri J, Shen C, Kazi DS, Yeh RW, Groeneveld PW. Association of Medicaid Expansion With Cardiovascular Mortality. JAMA Cardiol. 2019;4:671–679. doi: 10.1001/jamacardio.2019.1651

26. Khan SS, Matsushita K, Sang Y, Ballew SH, Grams ME, Surapaneni A, Blaha MJ, Carson AP, Chang AR, Ciemins E, et al. Development and Validation of the American Heart Association’s PREVENT Equations. Circulation. 2024;149:430–449. doi: 10.1161/CIRCULATIONAHA.123.067626

27. Centers for Disease Control and Prevention. Behavioral Risk Factor Surveillance System: Comparability of Data, BRFSS 2019. Atlanta, GA: US Department of Health and Human Services, Centers for Disease Control and Prevention; 2020. https://www.cdc.gov/brfss/annual_data/2019/pdf/compare-2019-508.pdf

28. Morgenstern H. Ecologic studies in epidemiology: concepts, principles, and methods. Annu Rev Public Health. 1995;16:61–81. doi: 10.1146/annurev.pu.16.050195.000425

29. Berrocal VJ, Gelfand AE, Holland DM. A Spatio-Temporal Downscaler for Output From Numerical Models. J Agric Biol Environ Stat. 2010;15:176–197. doi: 10.1007/s13253-009-0004-z

30. von Elm E, Altman DG, Egger M, Pocock SJ, Gotzsche PC, Vandenbroucke JP, STROBE Initiative. The Strengthening the Reporting of Observational Studies in Epidemiology (STROBE) statement: guidelines for reporting observational studies. J Clin Epidemiol. 2008;61:344–349. doi: 10.1016/j.jclinepi.2007.11.008

31. Benchimol EI, Smeeth L, Guttmann A, Harron K, Moher D, Petersen I, Sorensen HT, von Elm E, Langan SM, RECORD Working Committee. The REporting of studies Conducted using Observational Routinely-collected health Data (RECORD) statement. PLoS Med. 2015;12:e1001885. doi: 10.1371/journal.pmed.1001885

32. Ndumele CE, Rodriguez F, Dixon DL, Khan SS, Mukherjee D, Bajaj M, Bangalore S, Bozkurt B, Breathett K, Clarke SL, et al. 2026 AHA/ACC/ADA/ASN Guideline for the Prevention, Detection, Evaluation, and Management of Cardiovascular-Kidney-Metabolic Syndrome: A Report of the American College of Cardiology/American Heart Association Joint Committee on Clinical Practice Guidelines. Circulation. 2026;154:e50– e158. doi: 10.1161/CIR.0000000000001453

33. Centers for Disease Control and Prevention. Chronic Kidney Disease in the United States, 2023. Atlanta, GA: US Department of Health and Human Services, Centers for Disease Control and Prevention; 2023. https://www.cdc.gov/kidney-disease/php/data-research/index.html

34. U.S. Department of Agriculture, Economic Research Service. Food Access Research Atlas. https://www.ers.usda.gov/data-products/food-access-research-atlas. 2019. Accessed May 15, 2026.

35. U.S. Environmental Protection Agency. Air Quality System (AQS). https://www.epa.gov/aqs. 2020. Accessed May 15, 2026.

36. Bellander T, Berglind N, Gustavsson P, Jonson T, Nyberg F, Pershagen G, Jarup L. Using geographic information systems to assess individual historical exposure to air pollution from traffic and house heating in Stockholm. Environ Health Perspect. 2001;109:633– 639. doi: 10.1289/ehp.01109633

37. Cressie NAC. Statistics for spatial data. Rev. ed. New York: Wiley; 1993.

38. Riebler A, Sorbye SH, Simpson D, Rue H. An intuitive Bayesian spatial model for disease mapping that accounts for scaling. Stat Methods Med Res. 2016;25:1145–1165. doi: 10.1177/0962280216660421

39. Freni-Sterrantino A, Ventrucci M, Rue H. A note on intrinsic conditional autoregressive models for disconnected graphs. Spat Spatiotemporal Epidemiol. 2018;26:25–34. doi: 10.1016/j.sste.2018.04.002

40. Roberts DR, Bahn V, Ciuti S, Boyce MS, Elith J, Guillera-Arroita G, Hauenstein S, Lahoz-Monfort JJ, Schröder B, Thuiller W, et al. Cross-validation strategies for data with temporal, spatial, hierarchical, or phylogenetic structure. Ecography. 2017;40:913–929. doi: 10.1111/ecog.02881

41. Lundberg SM, Lee S-I. A Unified Approach to Interpreting Model Predictions. Advances in Neural Information Processing Systems. 2017;30.

42. Pebesma E. Simple Features for R: Standardized Support for Spatial Vector Data. The R Journal. 2018;10:439–446. doi: 10.32614/RJ-2018-009

43. Bivand RS, Wong DWS. Comparing implementations of global and local indicators of spatial association. TEST. 2018;27:716–748. doi: 10.1007/s11749-018-0599-x

44. Murakami D, Griffith DA. Spatially varying coefficient modeling for large datasets: Eliminating N from spatial regressions. Spatial Statistics. 2019;30:39–64. doi: 10.1016/j.spasta.2019.02.003

45. Rue H, Martino S, Chopin N. Approximate Bayesian Inference for Latent Gaussian models by using Integrated Nested Laplace Approximations. Journal of the Royal Statistical Society Series B: Statistical Methodology. 2009;71:319–392. doi: 10.1111/j.1467-9868.2008.00700.x

46. Chen T, Guestrin C. XGBoost: A Scalable Tree Boosting System. In: Proceedings of the 22nd ACM SIGKDD International Conference on Knowledge Discovery and Data Mining. San Francisco, California, USA: Association for Computing Machinery; 2016:785–794.

47. Mahoney M, Johnson L, Silge J, Frick H, Kuhn M, Beier C. Assessing the performance of spatial cross-validation approaches for models of spatially structured data. arXiv. Preprint. 2023. doi: 10.48550/arXiv.2303.07334

48. Stommel M, Schoenborn CA. Accuracy and usefulness of BMI measures based on self-reported weight and height: findings from the NHANES & NHIS 2001-2006. BMC Public Health. 2009;9:421. doi: 10.1186/1471-2458-9-421

49. Anselin L. Local Indicators of Spatial Association—LISA. Geographical Analysis. 1995;27:93–115. doi: 10.1111/j.1538-4632.1995.tb00338.x

50. Paciorek CJ. The importance of scale for spatial-confounding bias and precision of spatial regression estimators. Stat Sci. 2010;25:107–125. doi: 10.1214/10-STS326

51. Hodges JS, Reich BJ. Adding Spatially-Correlated Errors Can Mess Up the Fixed Effect You Love. The American Statistician. 2010;64:325–334. doi: 10.1198/tast.2010.10052

52. Ruiz JM, Steffen P, Smith TB. Hispanic mortality paradox: a systematic review and meta-analysis of the longitudinal literature. Am J Public Health. 2013;103:e52–60. doi: 10.2105/AJPH.2012.301103

53. Markides KS, Coreil J. The health of Hispanics in the southwestern United States: an epidemiologic paradox. Public Health Rep. 1986;101:253–265.

